# Combination of blood biomarkers and stroke scales improves identification of large vessel occlusions

**DOI:** 10.1101/2021.01.06.21249344

**Authors:** Edoardo Gaude, Barbara Nogueira, Sheila Graham, Sarah Smith, Lisa Shaw, Sara Graziadio, Gonzalo Ladreda Mochales, Marcos Ladreda Mochales, Philip Sloan, Joshua D. Bernstock, Shashank Shekhar, Toby I. Gropen, Christopher I. Price

## Abstract

**Background and Purpose:** Acute ischemic stroke caused by large vessel occlusions (LVO) is a major contributor to stroke deaths and disabilities; however, identification for emergency treatment is challenging.

**Aims:** To evaluate the diagnostic accuracy of a panel of biomarkers for LVO prediction.

**Methods:** 170 patients with suspected stroke were recruited retrospectively at one hospital. We analysed the plasma levels of D-dimer, OPN, OPG, GFAP, vWF, and ADAMTS13 in LVO vs non-LVO. Diagnostic performance was estimated by using blood biomarkers alone or in combination with NIHSS-derived stroke severity scales.

**Results:** Our patient cohort comprised 20% stroke mimics, 11% transient ischemic attack, 11% hemorrhagic stroke, 15% LVO ischemic stroke, 28% non-LVO ischemic stroke, and 15% ischemic stroke with unknown LVO status. Multivariable analysis found that the optimal set of blood biomarkers for LVO prediction was D-dimer (OR 15.4, 95% CI 4.9 to 57.6; p-value<0.001) and GFAP (OR 0.83, 95% CI 0.90 to 0.99; p-value=0.03). The combination of D-dimer and GFAP with stroke scales significantly improved LVO prediction, compared to the stroke scales alone (p-value<0.001). The combination of biomarkers with constructed FAST-ED or EMSA scales achieved an AUC of 95% (95% CI 91-100%) or 93% (CI 95% 89-97%), a sensitivity of 91% (95% CI 71-98%) or 86 (95% CI 66-97%), and a specificity of 95% (95% CI 89-98%) or 94% (95% CI 88-98%), for LVO prediction, respectively.

**Conclusions:** The combination of D-dimer, GFAP, and stroke scales could provide a simple and highly accurate tool for identifying LVO patients.

## Introduction

Stroke is among the leading causes of death and disability worldwide. Ischemic stroke caused by large vessel occlusion (LVO) contributes disproportionally to these figures and accounts for 62% of post-stroke disabilities and 96% of post-stroke mortality^1^.

Acute ischemic stroke patients with LVO can be effectively treated with endovascular thrombectomy (EVT)^2^, but this treatment is only available in comprehensive stroke centers (CSC) or other EVT-capable centers. Inter-hospital transfer of LVO patients from primary stroke centers to EVT-capable centers significantly delays time to treatment and leads to higher disability rates^3^. Identification of LVO patients in the pre-hospital setting (e.g. within an ambulance) could enable the transfer of LVO patients to EVT-capable centers directly, reducing time to treatment, functional disabilities, and deaths.

Multiple studies have investigated the ability of brief pre-hospital stroke assessment scales to identify LVO strokes in the field. Despite this, these severity scales lack the sensitivity and specificity required for triaging LVO patients with confidence, resulting in false negatives in patients with LVO and milder stroke as well as false positives in patients with stroke mimics or hemorrhagic strokes^4,5^. A more accurate diagnostic test able to complement these assessment scores and direct LVO patients to EVT-capable centers is much needed. Several research studies have investigated the ability of blood biomarkers to differentiate stroke from non-stroke patients, or ischemic from hemorrhagic stroke patients^6^. In addition, a number of studies have been dedicated to blood biomarkers of stroke etiology^7^. Yet, fewer studies have investigated the association of blood biomarker levels with LVO strokes specifically (i.e. as a subtype of ischemic stroke) and comprehensively (i.e. as a combination of different etiologic subtypes)^8,9^. Blood biomarkers for the identification of LVO strokes are yet to be identified and validated.

### Aims

The primary aim of this study was to identify the optimal combination of biomarkers that could differentiate between LVO and non-LVO. The screening panel of biomarkers comprised D-dimer, osteopontin (OPN) and osteoprotegerin (OPG), which have been associated with stroke etiologic subtypes^7,10^; glial fibrillary acidic protein (GFAP), an astrocyte marker found increased in hemorrhagic stroke^6^; von Willebrand factor (vWF), and a disintegrin and a metalloproteinase with a thrombospondin type I motif, member 13 (ADAMTS13), known markers of hemostasis that have been linked to ischemic stroke^11^. An additional analysis considered whether an optimal panel could increase the accuracy of stroke severity scales to differentiate between LVO and non-LVO stroke patients.

## Methods

### Study design and sample collection

This study was a retrospective observational study. Between August 2018 and February 2020, Cellular Pathology (CEPA) Biobank staff identified study eligible patients that had presented to the Emergency Department (ED) at the Royal Victoria Infirmary Hospital in Newcastle upon-Tyne (UK). Patients were identified based on the following criteria: 1) >18 years old; 2) evaluated in the ED for suspected stroke, as identified by ambulance paramedics, ED clinicians, and/or stroke specialist nurses; 3) <12 hours from their last known well or symptom onset time; 4) reperfusion therapy had not yet been received. As per clinical routine, whole venous blood was withdrawn at ED admission and stored at 4°C (protected from light). Whole blood diagnostic remnants of eligible patients were identified by biobank staff and centrifuged at 2000 x g for 15 minutes at 4°C and immediately frozen at −80°C. Frozen samples were then transferred in one batch to Pockit Diagnostics Ltd for biomarkers measurement. Ethics approval for this study was part of CEPA Biobank’s existing approval (NHS-HRA-North-East-Newcastle & North Tyneside 1 Research Ethics Committee, REC Reference: 17/NE/0070). All procedures were in accordance to institutional guidelines.

### Clinical data collection

The following routine clinical data were collected for eligible patients: demographics (age, gender), clinical characteristics (blood pressure, pulse, atrial fibrillation, hypertension), clinical laboratory results (hematology, biochemistry, blood lipids), admission NIHSS score, last known well or symptom onset time, blood withdrawal time, imaging findings within 1 hour, and final clinical diagnosis. Data were provided to Pockit diagnostics Ltd.

### Assigning a diagnostic category

Routine clinical data collected as described above were used to assign patients to the following diagnostic categories: LVO, non-LVO ischemic stroke, hemorrhagic stroke, transient ischemic attack (TIA), stroke mimic condition.

- Transient ischemic attack (TIA) and stroke mimic were assigned based on local specialist opinion
- Hemorrhagic stroke was assigned based on the presence of hemorrhage on brain imaging
- LVO required CT angiography confirmation by neuroradiologist report
- Remaining patients where a stroke specialist assigned a diagnosis of ischemic stroke were categorized as either non-LVO or not classifiable as follows:
  - non-LVO if CTA had been undertaken and LVO was not present OR if a CTA had not been undertaken but the NIHSS score on admission was <5. The latter was a pragmatic threshold reflecting little likelihood of eligibility for thrombectomy treatment^12^.
  - not classifiable if CTA had not been undertaken and NIHSS score on admission was >4.

Reference standard results were not available to the operators involved in blood biomarker measurements.

### Derivation of stroke scales from NIHSS score

FAST score was calculated by assigning 1 point for the presence of facial paresis (NIHSS item 4), 1 point for any arm weakness (NIHSS item 5a/b), and 1 point for any speech impairment (NIHSS item 9). FAST-ED was calculated as described by Lima et al^13^, RACE score was calculated as described by Perez de la Ossa^14^, C-STAT was calculated as described by Katz et al^15^, and EMSA was calculated as described by Gropen et al^16^.

### Measurement of blood biomarkers

Plasma biomarkers were measured using commercial enzyme-linked immunosorbent assays (ELISA) following manufacturer’s instructions. ELISA kits or matched antibody pairs were purchased from Abcam (Cambridge, UK): D-dimer (product number: ab196269), OPN (product number: ab100618), OPG (product number: ab100617), GFAP (product number: ab222279), vWF (product number: ab223864), and ADAMTS13 (product number: ab234559). Plasma sample dilutions for detecting each biomarker were as follows: D-dimer (1:80), OPN (1:2), OPG (1:6), GFAP (1:2), vWF (1:4000), ADAMTS13 (1:800). All samples were analysed in duplicate and the mean value was used for quantification. All readings were performed with a Multiskan™ FC spectrophotometer (Thermo-Fisher Scientific, Catalog Number 51119000). For all biomarkers, the average coefficient of variation was <10%. GraphPad Prism version 8.4.3 was used for biomarker quantification.

### Statistical analysis

The proposed intended use of the blood biomarkers is to rule in patients with LVO to redirect patients to EVT-capable centres from a population of suspected stroke, thus we powered the study on specificity. Assuming a prevalence of 15%, a minimum specificity of 90%, a two-tailed 5% type I error rate (α) and 90% power (β), a sample size of 161 suspected patients with 25 LVO would detect a specificity of 97% (95%CI: 90%–100%) for LVO.

Symptom onset to blood collection time (OBT) was calculated from last known well/onset time and time of blood withdrawal. Where patients were reported to have woken up with symptoms, the time since last known well was assumed to be midnight of the previous day. Where a range for onset time/last known well time was provided, the first time value was used to calculate OBT.

To compare the levels of blood biomarkers and clinical variables in LVO and non-LVO patients, distribution normality of continuous variables was assessed by Shapiro–Wilk test and: i) for normally distributed variables, Welch’s t-test and mean ± standard deviation (SD) were used or ii) for non-normally distributed variables Wilcoxon-Mann–Whitney U test, median and interquartile range (IQR) were used. Categorical variables were assessed by Pearson’s chi-square test. To compare variables between subtypes of suspected stroke, ANOVA was used. When >10 variables were tested at the same time, multiple hypothesis correction was performed using Benjamini-Hochberg’s method. Analysis of variance was used to assess the interaction between all blood biomarkers and subtypes of suspected stroke. If overall significance was confirmed, pairwise comparisons were performed with Tukey’s test.

To identify the optimal panel of blood biomarkers for LVO prediction, we used a multivariate logistic regression with Diagnosis (LVO vs non-LVO) as outcome variable and plasma levels of D-dimer, GFAP, OPN, OPG, vWF, and ADAMTS13 as exploratory variables.

Bidirectional stepwise elimination based on Akaike information criterion (AIC) levels was used for model selection. Linearity between predictors and outcome measure was assessed through logarithmic and quadratic transformation. Transformations were selected based on the AIC.

To investigate whether the addition of blood biomarkers improved the accuracy of stroke severity scales for LVO identification, we used a second multivariate logistic with Diagnosis as the outcome variable and the optimal panel and one of the stroke severity scales (FAST, FAST-ED, RACE, C-STAT, or EMSA) as exploratory variables. We used this approach because the scales were highly correlated, and, since a comparison of different severity scales was outside the scope of this work, this approach reduced the level of collinearity in the model.

To assess the goodness of fit of the blood biomarker panel and the stroke scales, the likelihood ratio test (LR) and AIC were used. The area under the receiver operating characteristic curve (AUC) with 95% CIs was used as a measure of discrimination. At selected cut-off points sensitivity, specificity, positive likelihood ratio (LR+), and negative likelihood ratio (LR-) were also evaluated. For each model, the cut-off point was selected by maximising specificity for LVO prediction while maintaining a minimum specificity level of 90%, in line with our power analysis. Correction for optimistic predictions was performed through bootstrapping with 2000 resamples, and presented with confidence intervals (CI). All analyses were performed with R version 3.6.2 with the help of RStudio version 1.2.5033 by using the packages nnet, ROCR, caret, tidyverse, oddsratio, lmtest, and OptimalCutpoints.

## Results

### 1.1. Study enrolment

Data were obtained from 170 patients with suspected stroke. Blood samples from 19 patients were utilized for immunoassay testing and then excluded from further use. Data from a further 23 patients could not be used because they had ischemic stroke which could not be categorized as LVO or non LVO. The remaining 128 patients was 21% lower than our sample size estimation, due to disruptions caused by the COVID19 pandemic. The final cohort of 128 suspected stroke patients categorized as follows (supplementary figure 1): LVO ischemic stroke (n=23, 18%), non-LVO ischemic stroke (n=42, 33%), hemorrhagic stroke (n=16, 12.5%), stroke mimic (n=31, 24%) and transient ischemic attack (n=16, 12.5%). Stroke mimic diagnoses were as follows: anaemia (3%), anxiety (10%), Bell’s palsy (7%), delirium (3%), dementia (3%), depression (3%), dysphasia (3%), metastatic cancer (3%), migraine (16%), seizure (13%), syncope (13%), vertigo (13%), and undetermined (10%).

### 1.2. Patient characteristics

Clinical characteristics for LVO and all non-LVO patients are reported in Table 1. We found significant differences between LVO and all non-LVO in NIHSS score (18±9 and 3±5, p-value<0.001), presence of atrial fibrillation (52% and 10%, p-value<0.001), and systolic blood pressure (140±22 and 157±29 mmHg, p-value=0.03). Of note, age, gender, OBT, or blood processing time were not different between LVO and non-LVO.

**Table 1.**
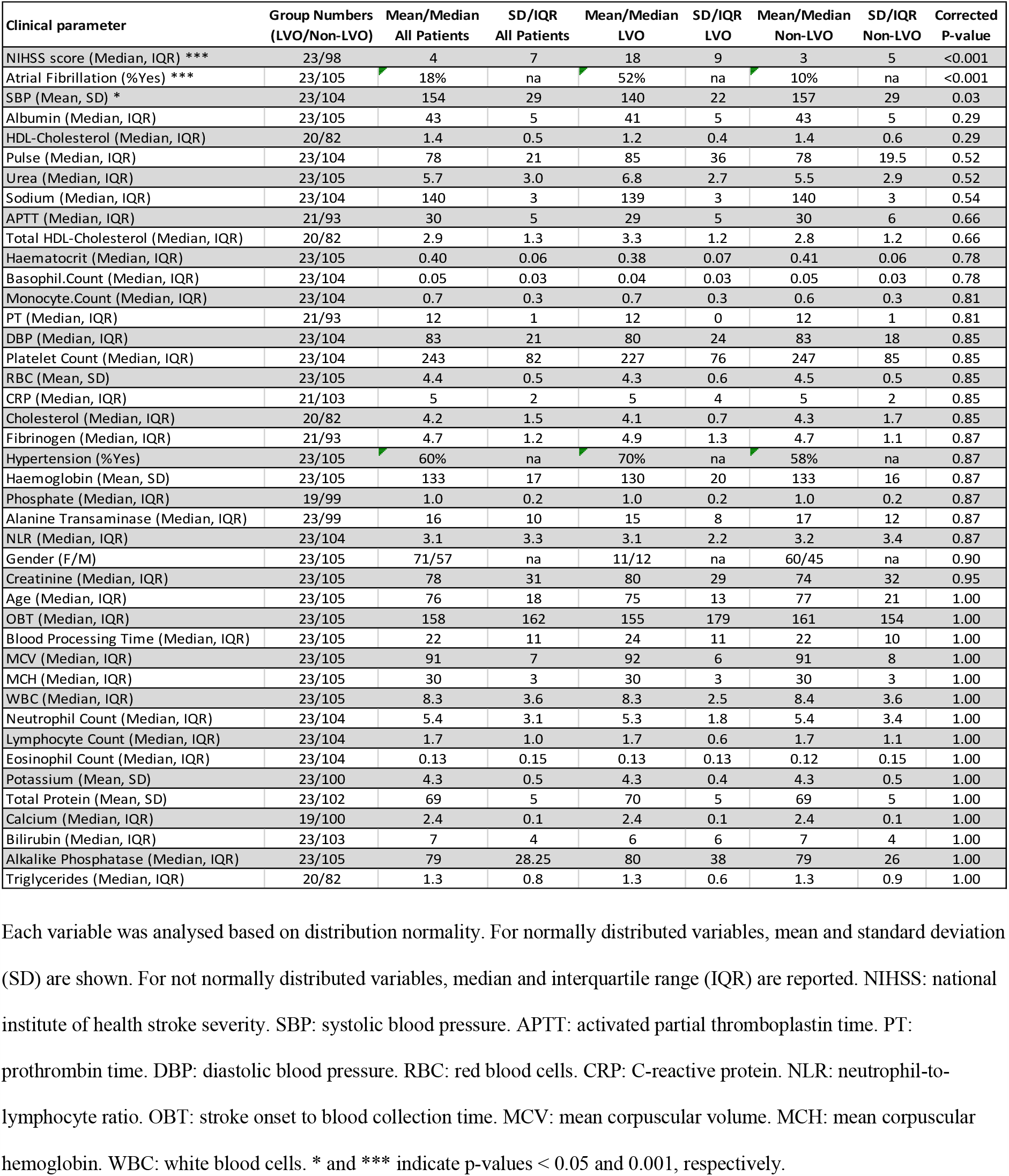
Univariate analysis of 44 clinical variables in LVO and non-LVO stroke patients.

The levels of D-dimer, OPN, OPG, vWF, ADAMTS13, and GFAP in the plasma of LVO and non-LVO patients are reported in Supplementary Table 1. We found statistically significant differences between LVO and all non-LVO for the following blood biomarkers: D-dimer (1.3±2.0 and 0.4±0.4 µg/mL, p-value<0.001); OPN (1.7±1.1 and 1.2±1.1 ng/mL, p-value=0.02); OPG (125±60 and 96±54 pg/mL, p-value=0.01). We found significant interaction between blood biomarkers and different subtypes of suspected stroke: D-dimer, OPN, and GFAP were significantly associated with specific patient subgroups (Supplementary Table 2).

### 1.3. Optimal blood biomarker panel for LVO prediction

Levels of D-dimer, OPN, OPG, and ADAMTS13 were log transformed, while levels of vWF underwent quadratic transformation; no transformation was applied to GFAP levels. Potentially useful predictors with univariate analysis for LVO accuracy were D-dimer (OR 1.13, 95% CI 1.07-1.20; p-value<0.001), and OPG (OR 1.10, 95% CI 1.01 to 1.22; p-value=0.04; Table 2). The optimal set of blood biomarkers for LVO prediction identified by the logistic regression was D-dimer (OR 15.4, 95% CI 4.9-57.6; p-value<0.0011) and GFAP (OR 0.83, 95% CI 0.50-0.99; p-value=0.03; Table 3). The AUC of the model with D-dimer and GFAP for LVO prediction was 81% (95% CI 74-88%; Table 4). The cut-off that maximized specificity was 0.33 and gave a sensitivity of 56% (95% CI 34-77%) at a specificity of 92% (95% CI 85-96%). LR+ was 7 (95% CI 3-15), and LR-0.47 (95% CI 0.3-0.76; Table 5).

**Table 2.** Univariate logistic regression for LVO prediction.

|  | Beta | CI | OR | Pvalue |
| --- | --- | --- | --- | --- |
| DDimer | 2.898308631 | 0.6811 to 1.8216 | 1.129 (1.07 to 1.2) | 2.00E-05 |
| OPN | 1.036734324 | -0.0559 to 1.2219 | 1.058 (0.994 to 1.13) | 0.07773 |
| OPG | 1.25089079 | 0.0743 to 1.9804 | 1.102 (1.007 to 1.219) | 0.04088 |
| GFAP | -4.176985961 | -0.0096 to 2e-04 | 1 (0.999 to 1) | 0.48998 |
| vWF | 0.80427253 | -2e-04 to 9e-04 | 1 (1 to 1) | 0.18587 |
| ADAMTS13 | 0.417022833 | -1.1465 to 2.5246 | 1.065 (0.892 to 1.287) | 0.50088 |

The p-values indicate statistical significance of the covariate in the model.

**Table 3.** Univariate and multivariable logistic regression models for LVO prediction.

| Model | Parameters | Beta | CI | OR (95% CI) | Pvalue |
| --- | --- | --- | --- | --- | --- |
| Biomarkers | D-dimer | 3.3 | 0.8 to 2.1 | 16 (5-60) | <0.001 |
|  | GFAP | -1.6 | -1.5 to -0.04 | 0.3 (0.06-0.9) | 0.03 |
| C-STAT | C-STAT | 2.3 | 0.5 to 1.3 | 2.4 (1.6-3.6) | <0.001 |
| C-STAT + biomarkers | C-STAT | 3.0 | 0.6 to 1.7 | 3 (1.8-5.7) | <0.001 |
|  | D-dimer | 3.3 | 0.8 to 2.2 | 16.6 (4.5-76) | <0.001 |
|  | GFAP | -2.5 | -1.8 to -0.36 | 0.15 (0.03-0.5) | <0.01 |
| EMSA | EMSA | 4.5 | 0.56 to 1.4 | 2.5 (1.7-4.1) | <0.001 |
| EMSA + biomarkers | EMSA | 5.7 | 0.7 to 1.9 | 3.3 (2-6.4) | <0.001 |
|  | D-dimer | 3.4 | 0.7 to 2.3 | 18.4 (4.4-104) | <0.001 |
|  | GFAP | -3.2 | -2.3 to -0.4 | 0.09 (0.01-0.44) | 0.01 |
| FAST | FAST | 3.8 | 0.9 to 2.3 | 4.6 (2.5-9.7) | <0.001 |
| FAST + biomarkers | FAST | 4.9 | 1.14 to 3 | 7.1 (3.1-20) | <0.001 |
|  | D-dimer | 3.5 | 0.8 to 2.4 | 20 (5-112) | <0.001 |
|  | GFAP | -2.8 | -2.1 to -0.35 | 0.12 (0.02-0.5) | 0.01 |
| FAST-ED | FAST-ED | 4.1 | 0.48 to 1.03 | 2.07 (1.6-2.8) | <0.001 |
| FAST-ED + biomarkers | FAST-ED | 6.3 | 0.7 to 1.7 | 3 (2-5.3) | <0.001 |
|  | D-dimer | 3.66 | 0.7 to 2.7 | 23 (4-210) | 0.001 |
|  | GFAP | -3.95 | -2.8 to -0.7 | 0.05 (0-0.23) | <0.01 |
| RACE | RACE | 3.9 | 0.36 to 0.74 | 1.7 (1.43-2.1) | <0.001 |
| RACE + biomarkers | RACE | 5.01 | 0.43 to 1.05 | 2 (1.5-2.9) | <0.001 |
|  | D-dimer | 3.5 | 0.7 to 2.5 | 20.4 (4.1-138) | <0.001 |
|  | GFAP | -3.57 | -2.9 to -0.43 | 0.06 (0-0.43) | 0.03 |

The p-values indicate statistical significance of the model covariate.

**Table 4.** Model comparisons.

|  | **AIC** | **Deviance** | **AUC** | **LR (df), p-value** |
| --- | --- | --- | --- | --- |
| D-dimer+GFAP | 100 | 94 | 81 (74-88) | - |
| FAST | 93 | 89 | 83 (78-88) | - |
| FAST+D-dimer+GFAP | 72 | 64 | 93 (90-97) | 26 (4), <0.001 |
| FAST-ED | 78 | 74 | 91 (86-95) | - |
| FAST-ED+D-dimer+GFAP | 51 | 43 | 95 (91-100) | 31 (4), <0.001 |
| RACE | 79 | 75 | 87 (82-93) | - |
| RACE+D-dimer+GFAP | 59 | 51 | 93 (89-98) | 24 (4), <0.001 |
| C-STAT | 103 | 99 | 79 (72-86) | - |
| C-STAT+D-dimer+GFAP | 79 | 71 | 88 (81-94) | 27 (4), <0.001 |
| EMSA | 90 | 86 | 84 (79-89) | - |
| EMSA+D-dimer+GFAP | 70 | 62 | 93 (89-97) | 23 (4), <0.001 |

LR p-values refer to the comparison between the model with a stroke scale alone and the corresponding model with the addition of biomarkers.

**Table 5.** Internal validation of diagnostic accuracy.

|  | **Cut-off** | **Sensitivity** | **Specificity** | **LR+** | **LR-** |
| --- | --- | --- | --- | --- | --- |
| D-dimer+GFAP | 0.33 | 57 (34-77) | 92 (85-96) | 7 (3-15) | 0.47 (0.3-0.76) |
| C-STAT | 0.4 | 39 (20-61) | 93 (86-97) | 6 (2-13) | 0.66 (0.47-0.91) |
| C-STAT+D-dimer+GFAP | 0.39 | 74 (52-90) | 93 (86-97) | 10 (5-22) | 0.28 (0.14-0.56) |
| EMSA | 0.66 | 26 (10-48) | 98 (93-100) | 13 (3-60) | 0.75 (0.59-0.96) |
| EMSA+D-dimer+GFAP | 0.34 | 87 (66-97) | 95 (89-98) | 17 (7-41) | 0.14 (0.05-0.39) |
| FAST | 0.62 | 35 (16-57) | 92 (85-96) | 4 (2-10) | 0.71 (0.52-0.96) |
| FAST+D-dimer+GFAP | 0.33 | 78 (56-93) | 93 (86-97) | 11 (5-23) | 0.23 (0.11-0.51) |
| FAST-ED | 0.37 | 74 (52-90) | 92 (85-96) | 9 (5-19) | 0.28 (0.14-0.57) |
| FAST-ED+D-dimer+GFAP | 0.39 | 91 (72-99) | 96 (90-99) | 23 (9-60) | 0.09 (0.02-0.34) |
| RACE | 0.38 | 70 (47-87) | 93 (86-97) | 10 (5-21) | 0.33 (0.18-0.61) |
| RACE+D-dimer+GFAP | 0.24 | 83 (61-95) | 93 (86-97) | 12 (6-24) | 0.19 (0.08-0.46) |

### 1.4. Combination of blood biomarkers and stroke scales

We report stroke severity scales in LVO and non-LVO patients in Supplementary Table 3. The AUCs obtained from the univariate logistic regressions of each stroke scale for LVO discrimination were (Table 4): C-STAT 79% (95% CI 72-86%), EMSA 84% (95% CI 79-89%), FAST 83% (95% CI 78-88%), FAST-ED 91% (95% CI 86-95%), RACE 87% (95% CI 82-93%).

The addition of the optimal biomarker panel previously identified (D-dimer and GFAP) to the stroke severity models resulted in models with a lower AIC, a higher AUC and a significant LR test (Table 4 and Figure 1). In line with this, D-dimer and GFAP substantially improved sensitivity without adversely affecting specificity of clinical scales, such that all achieved high performance for LVO prediction (Table 5); of note, addition of D-dimer and GFAP to FAST-ED or EMSA resulted in the highest LR+ of 23 (95% CI 9-60) or 17 (95% CI 7-41), LR-of 0.09 (95% CI 0.02-0.34) or 0.14 (95% CI 0.05-0.39), sensitivity of 91% (95% CI 72-99%) or 87 (95% CI 66-97%), and specificity of 96% (95% CI 90-99%) or 95% (95% CI 89-98%), respectively. Overall, the addition of D-dimer and GFAP to stroke scales allowed to increase the number of true positives, while reducing the number of false positives due to hemorrhage or mimics (Supplementary Table 4), depending on the scale used.

**Figure 1.**
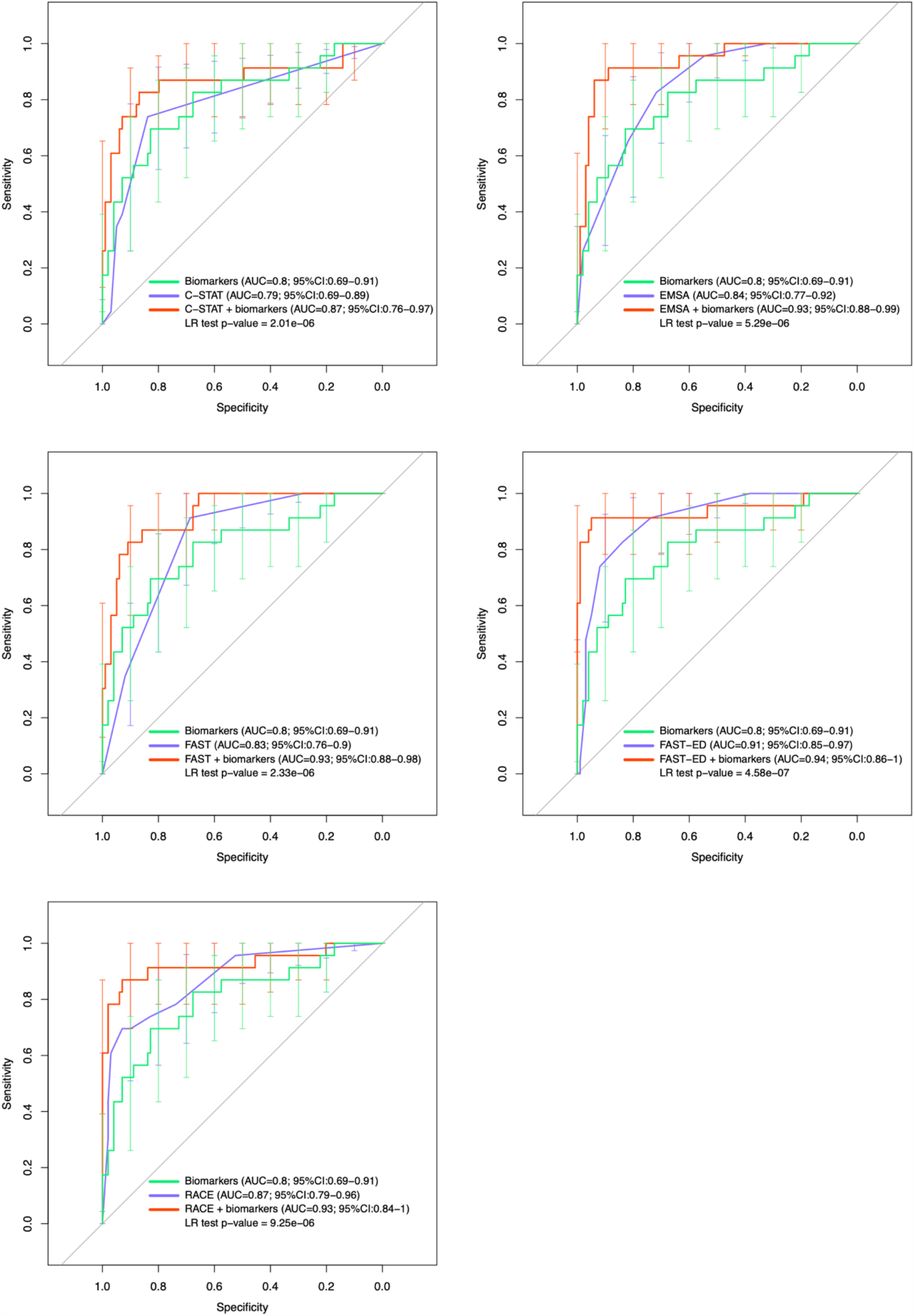
ROC curves of biomarker model and stroke scales models with or without the addition of biomarkers. Error bars indicate sensitivity confidence intervals. AUC with 95% CIs are shown for each model.

## Discussion

This study demonstrates that a biomarker panel composed of D-dimer and GFAP could provide a valuable tool for highly specific identification of stroke patients with LVO. Moreover, we have shown that combining D-dimer and GFAP with pre-hospital stroke scales can result in high predictive power (AUC=95%) for LVO in a hospital population of patients with suspected stroke.

D-dimer has been previously associated with cardioembolic strokes^17,18^, but few reports have investigated its usefulness for LVO identification. Occlusions in the main stem of intracranial arteries or in the cervical arteries have been linked to increased levels of plasma D-dimer^18^. In a separate study, cut-off values of D-dimer at 445 ng/mL or 300 ng/mL (depending on detection technique) distinguished large artery from small artery strokes^17^. Nevertheless, relevant limitations of these studies are the lack of CTA results for confirmation of LVO and absence of stroke mimics within the study population. While previously published studies focussed on comparing D-dimer levels between subtypes of ischemic stroke or between ischemic stroke and control subjects, our study is the first to assess D-dimer levels in LVO patients compared to a heterogeneous group of suspected stroke patients encompassing hemorrhagic stroke, non-LVO ischemic stroke, stroke mimics, and TIAs. The population recruited in our study more accurately reflects the population that is present in an ambulance setting or in the ED.

Previous work has shown that GFAP levels are elevated in hemorrhagic stroke patients, as compared to ischemic strokes, stroke mimics and/or TIAs^19,20^. Our findings confirm this evidence, showing significantly higher plasma levels of GFAP in hemorrhagic patients. To our knowledge, no studies have addressed directly the role of GFAP in the identification of LVO patients. Our study demonstrates that, when measured with D-dimer, GFAP can significantly improve LVO identification, by ruling out hemorrhagic patients from the population of suspected strokes.

Few studies have sought to examine the diagnostic performance of individual vs combinations of biomarkers for stroke identification. For example, Montaner et al measured a panel of ten different blood proteins between the etiologic subtypes of ischemic stroke and found that brain natriuretic peptide (BNP), D-dimer, and soluble receptor of advanced glycation end products (RAGE) were increased in ischemic stroke of cardioembolic origin^7^. Notably, prediction of cardioembolic stroke was poorer when BNP or D-dimer were used in isolation, while diagnostic performance increased when both biomarkers were combined to categorize stroke subtypes^7^. In line with this evidence, we showed that the combination of multiple biomarkers into a panel permits higher diagnostic performance for LVO identification, compared to the use of individual biomarkers.

We estimated the diagnostic performance of validated stroke severity scales for LVO prediction and observed that their overall accuracy was higher, compared to previous studies^13–16,21,22^. This could be due to derivation of stroke scales from the NIHSS score, which was taken by ED clinicians. This may be particularly relevant for the FAST-ED and RACE which were found to be more complex compared to the FAST or EMSA^16^. Other studies have showed that the combination of clinical variables with blood biomarkers can improve stroke diagnosis. Brouns and colleagues reported that the addition of the Oxford Community Stroke Project (OCSP) classification to D-dimer measurements could improve the accuracy of lacunar stroke identification from 88% to 98%^25^. Montaner et al showed that a logistic model built on the levels of D-dimer and BNP, together with the NIHSS score and other clinical variables, resulted in better diagnostic accuracy for cardioembolism, compared to the biomarkers alone^7^. In line with this evidence, here we demonstrate that a model built on D-dimer and GFAP, in conjunction with pre-hospital stroke scales, can lead to higher predictive ability for LVO, compared to the use of blood biomarkers, or stroke scales, in isolation. Each of the NIHSS-derived scales had advantages and disadvantages; the combination of the biomarkers with FAST-ED resulted in the highest diagnostic performance for LVO overall. Nevertheless, the collection of FAST-ED is more complex compared to other stroke scales, such as FAST or EMSA^16^, which may be preferred for field applications. We showed that the combination of D-dimer and GFAP with FAST or EMSA achieved sufficient LVO prediction (AUC=93%), assuming these results are replicated in further studies. The combination of D-dimer and GFAP with FAST or EMSA may be easier to replicate in the prehospital setting.

High levels of specificity (or positive predictive value) would be required to identify LVO patients in the field, in order to deviate the ambulance journey from transportation to the nearest stroke centre, towards the nearest EVT-capable centre^4^. Our findings indicate that combining our biomarkers with any of a number of pre-hospital stroke scales has the potential to provide the level of diagnostic performance needed to triage LVO patients with reasonable confidence.

Our study had several practical limitations. We used blood samples derived from routine clinical tests that were stored in the dark at +4° before processing. Although previous studies have shown stability of some of the tested blood biomarkers up to 240h at +4°C^23,24^, we cannot exclude that some of our negative results could be due to protein degradation. Biomarker measurement was performed with standard laboratory immunoassays which are inherently variable and therefore would need comparison and validation in a future study. Symptom onset times were obtained from medical records by non specialists and there may be some inaccuracy but it is likely that patients were within 12 hours of onset. Categorisation into LVO and non-LVO patients was based on routinely available clinical and imaging findings, including a pragmatic decision for handling ischemic stroke patients where CTA was not available. This pragmatic method may have led to loss of LVO cases from the cohort (excluded as no CTA) or incorrect assignment of mild stroke as non-LVO (ischemic stroke with NIHSS < 5 assigned as non-LVO). A further study should include CTA for all ischemic stroke patients and independent adjudication of LVO. Stroke scales were derived based on hospital-completed NIHSS scores, which may not reflect the results that would be obtained by paramedics completion of these scales in the field.. We recruited less patients than the sample size calculation which may have reduced the confidence in estimates of diagnostic performance for LVO identification.

In conclusion, our study suggests that combining the measurement of plasma D-dimer and GFAP with stroke severity scales may offer a valuable tool for early identification of LVO stroke patients. These findings need to be validated in a prospective study and if the tool is to be used in the pre-hospital setting, a point-of-care device is required. Evaluation of cost-effectiveness and clinical utility of the tool would also be needed before introduction into clinical practice.

## Data Availability

The data that support the findings of this study, as well as the study protocol, are available from the corresponding author upon reasonable request.

## Acknowledgements

E.G., G.L.M., M.L.M., C.I.P., L.S., S. Graziadio, S. Shekar, and T.I.G. contributed to study ideation and design. S. Graham and S. Smith performed sample recruitment, blood processing, and collected all clinical data. E.G. and B.N. performed all biomarkers measurements. E.G. performed all statistical analyses with advice from S. Graziadio. All authors contributed to writing this manuscript.

