## Supplemental Material for "Combination of blood biomarkers and stroke scales improves identification of large vessel occlusions"

### Supplemental Tables

Supplemental Table 1.

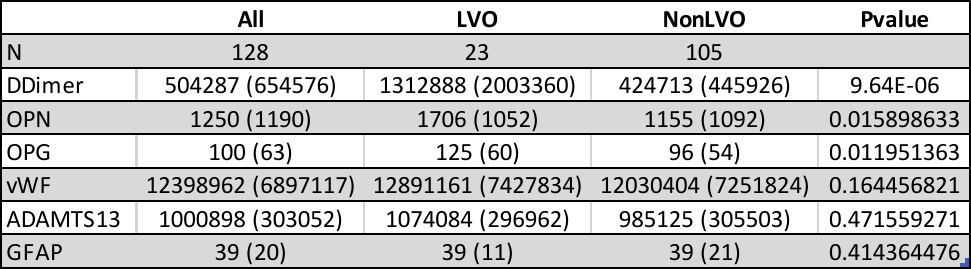

Plasma concentrations of blood biomarkers measured in all patients, LVO, and non-LVO patients. Median values are reported with IQR values shown in brackets. Wilcoxon-Mann Whitney p-values are shown.

Supplemental Table 2.

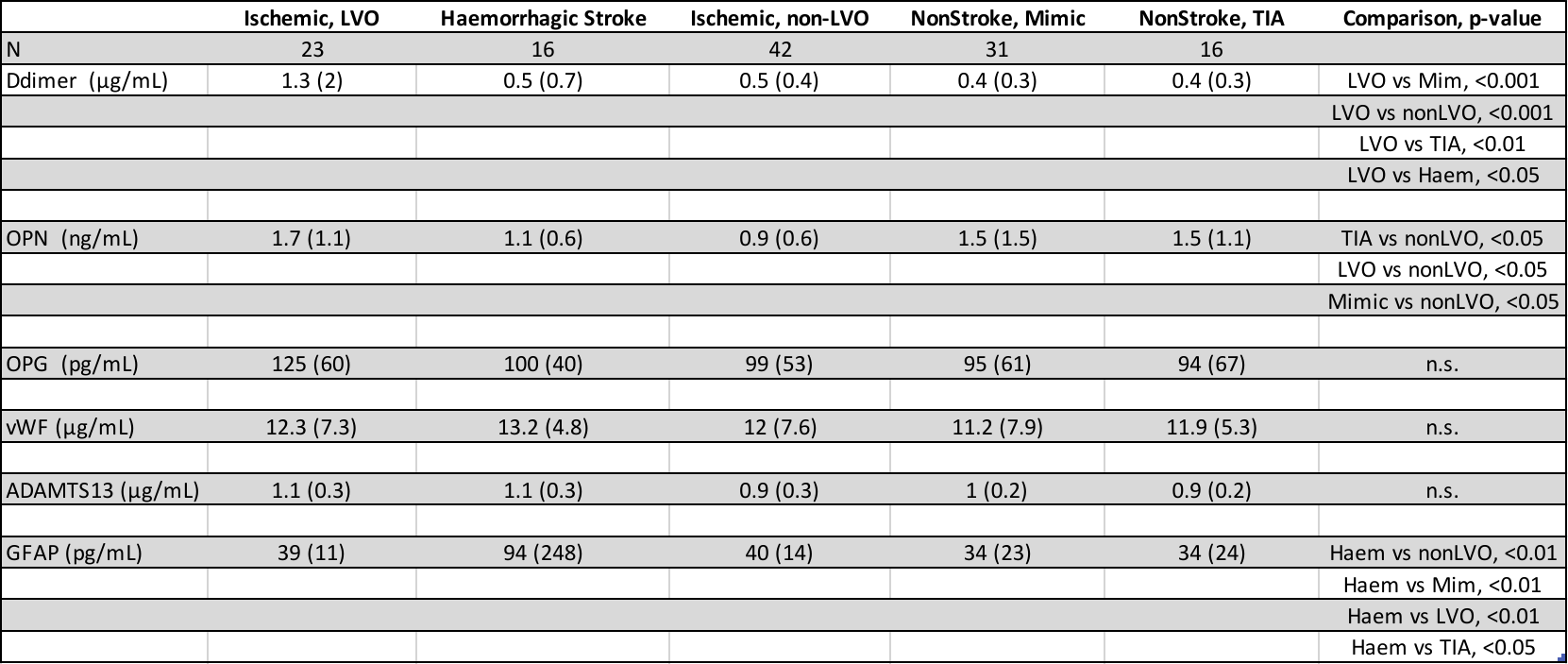

Plasma concentrations of blood biomarkers measured in different subtypes of suspected stroke patients. Median values are reported (pg/mL), with IQR values shown in brackets. For significant associations, Tukey’s p-values are reported next to the corresponding pairwise comparison.

Supplemental Table 3.

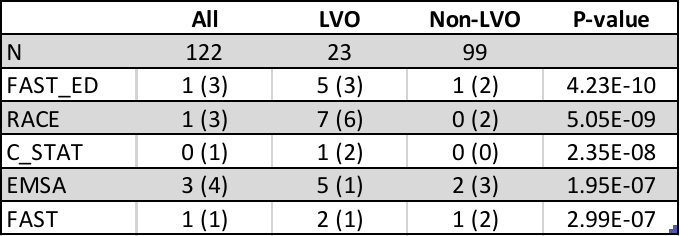

Median values of stroke severity scales obtained for all patients, LVO, and non-LVO patients. IQR values shown in brackets. Wilcoxon-Mann Whitney was used to calculate p-values.

Supplemental Table 4.

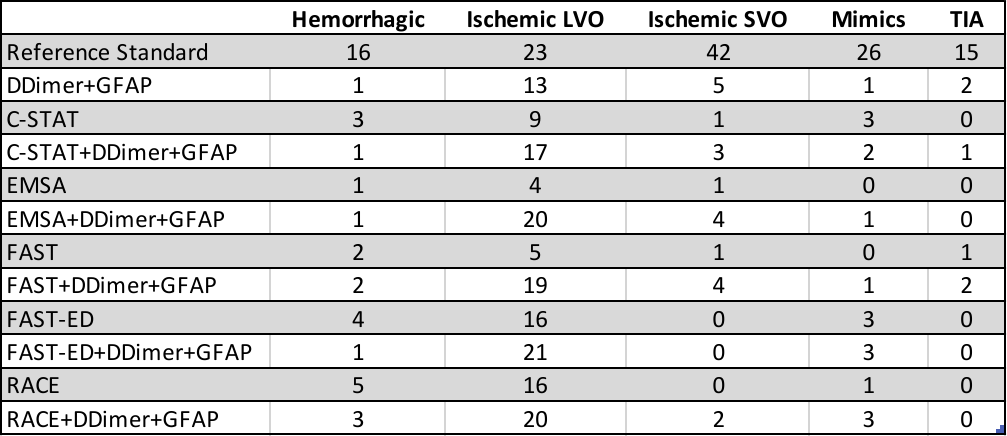

Number of patients selected by each model or stroke scale, compared to the reference standard. Number of patients selected are divided into the different subtypes of suspected stroke.

### Supplemental Figures

Supplemental Figure 1. Diagnostic categorization.

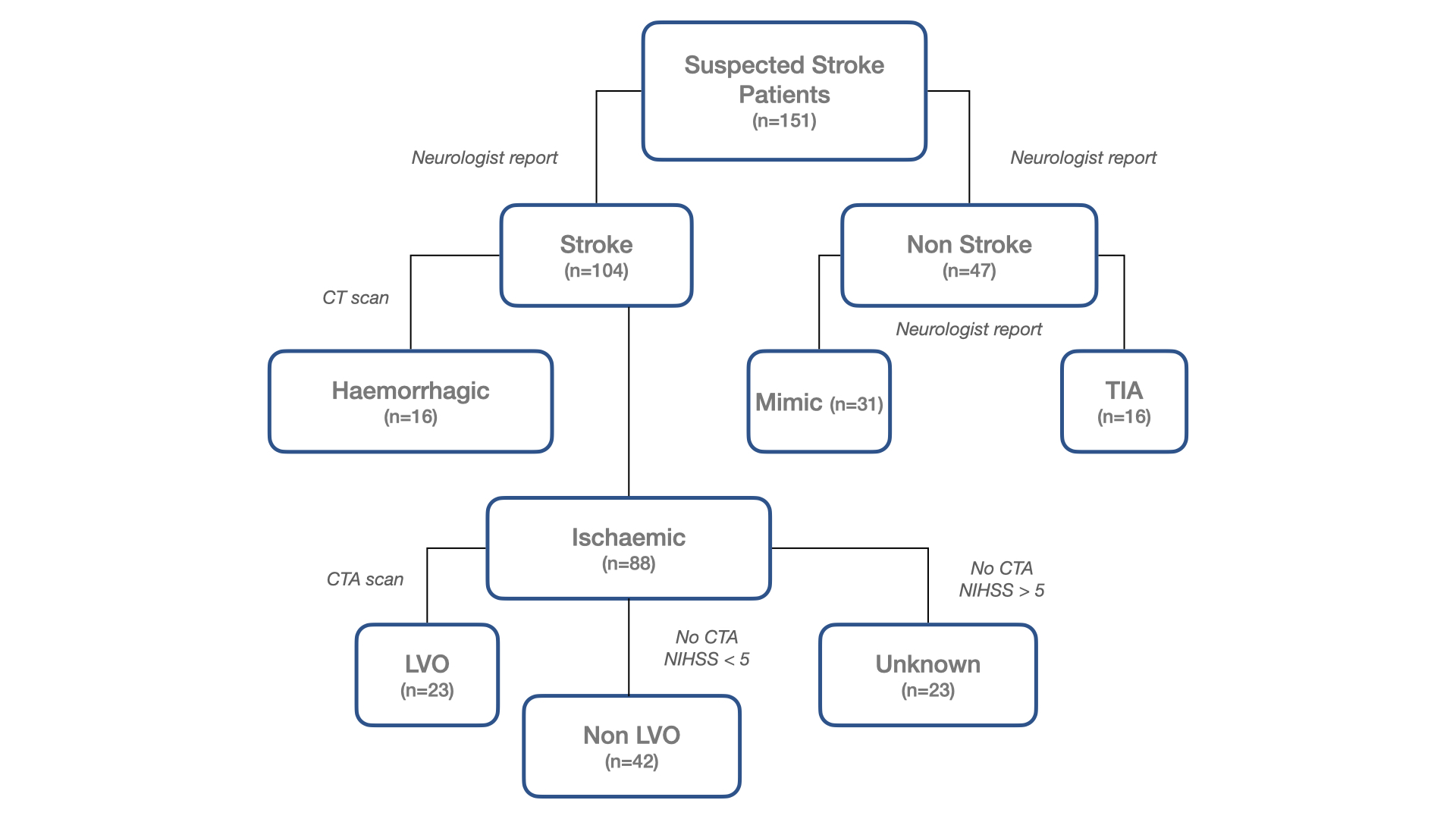

### Supplemental Figure Legends

Supplemental Figure 1. Each box represents different types of stroke subtype or suspected stroke. Number of each patient categorised in different subtypes is reported in brackets. Italic text outside each box indicates the methods used to assign diagnosis.
